# REDUCING DOOR‐TO‐BALLOON TIME USING EMS INITIATED APP BASED COMMUNICATION

**DOI:** 10.1101/2023.07.06.23292332

**Authors:** Christian Abrahim, Alina Capatina, Arvind Kalyan‐Sundaram, Amir Lotfi

## Abstract

**Background:** Reducing door-to-balloon (D2B) time for ST-Segment Elevation Myocardial Infarctions (STEMI) has been shown to be effective in improving outcomes. Delays still occur due to various factors such as time to lab activation and diagnostic clarification in equivocal cases. We propose that early communication through a mobile application between Emergency Medical Services (EMS) and in-hospital providers can reduce EMS-to-balloon time and D2B time.

**Methods:** We implemented the General Devices Company, Inc. e-BridgeTM mobile application for EMS providers. The app allowed real-time communication and data transmission between EMS providers and the cardiac catheterization lab and emergency department staff. A single-center, retrospective observational study was conducted on 795 STEMI activations undergoing emergent coronary angiography between January 2017 and July 2020. After exclusions, sample size was determined to be 428. The implementation of the app occurred on May 14th, 2019, and EMS transmissions were utilized in 132 patient cases during the study period.

**Results:** The implementation of the application resulted in a significant reduction in D2B time by 7.2 minutes (61.2 minutes vs. 68.36 minutes, p=0.005, CI -12.2 to -2.14) and ED board time by 4.41 minutes (29.38 minutes vs. 24.98 minutes, p=0.01, CI -7.8 to -1). There was a trend towards a reduction in First Medical Contact to balloon time, although it was not statistically significant. Further analysis revealed that the application did not significantly reduce D2B time or First Medical Contact to balloon time during the day shift but had a significant improvement in these metrics during the night shift.

**Conclusion:** The implementation of app-based communication between EMS providers and in-hospital providers using GPS tracking resulted in statistically significant reductions in First Medical Contact to balloon time and D2B time at our institution. This approach holds promise in improving the timely management of STEMI patients, particularly during the night shift.

## Introduction

The door-to-balloon (DTB) time is a well-established benchmark for measuring the quality of percutaneous primary coronary intervention in patients with ST-elevation myocardial infarction (STEMI). An incremental delay in DTB time is associated with an increased relative risk of in-hospital mortality and worse patient outcomes. Early activation of the cardiac catheterization lab by emergency medical services (EMS) has been shown to be an effective strategy in reducing DTB time. Communication between EMS and in-hospital providers plays a crucial role in facilitating appropriate triage, diagnosis, and initial management.

New communication technologies, such as real-time wireless transmission of data from the field to receiving hospitals, aim to minimize delays in diagnosis and triage before patients arrive at the hospital. While studies have shown the benefits of app-based communication for stroke patients, the impact on DTB times for STEMI patients has been inconclusive. However, a single-center study in Beijing demonstrated that a telemedicine app called the Tiantanzhixin app improved DTB times both before and after the COVID-19 pandemic. This app facilitated real-time communication and online consultation between patients and the hospital.

In an effort to enhance prehospital communication and transmit electrocardiograms (EKGs) between EMS, the emergency department (ED), and the STEMI team, we acquired the General Devices Company, Inc. e-BridgeTM mobile telemedicine application. We provided training to all EMS agencies free of cost with the primary objective of improving the time from first medical contact to balloon and door-to-balloon time. As of now, no similar studies have been published, making this endeavor an innovative approach to improving STEMI patient care.

## Methods

The project was implemented following comprehensive training provided to participating EMS agencies, ED staff, cath lab staff, and physicians. User accounts were granted to private EMS agencies and hospital personnel at no cost. These accounts are password protected and adhere to HIPAA security standards, allowing for secure usage on any smart device. In addition, smart tablets were strategically installed throughout the ED and cath lab areas.

To conduct our study, we performed a Retrospective Observational Cohort Study, which received review and approval for waiver from the Baystate Medical Center Institutional Review Board. The study population comprised individuals aged 18 years or older with ST-segment elevation of more than 1 mm in 2 contiguous ECG leads lasting for more than 30 minutes, or those with new left bundle branch block or a clinical syndrome indicative of acute evolving transmural myocardial infarction (MI) necessitating immediate interventional reperfusion therapy. We excluded cases involving interfacility transfers, STEMI diagnoses based on subsequent ECGs, late presentation of STEMI, known bleeding disorders, cases where no PCI procedure was performed, and instances where thrombolytics were administered. Cases with delays in PCI due to non-systemic factors were not excluded in either group. The primary endpoints assessed in this study included first medical contact to balloon time, door to balloon time, door to leave ED time, and EMS travel time in minutes. Additionally, we collected data on the rate of false STEMI activations.

To protect patient confidentiality, all data was de-identified and stored on a password-protected computer system. Patient anonymity was ensured by coding all collected data. Importantly, none of the investigators involved in this study had any personal or financial interests that could potentially influence their objectivity in data gathering, analysis, or the recommendations they made based on the study results.

For the statistical analysis of the collected data, we utilized the SPSS application.

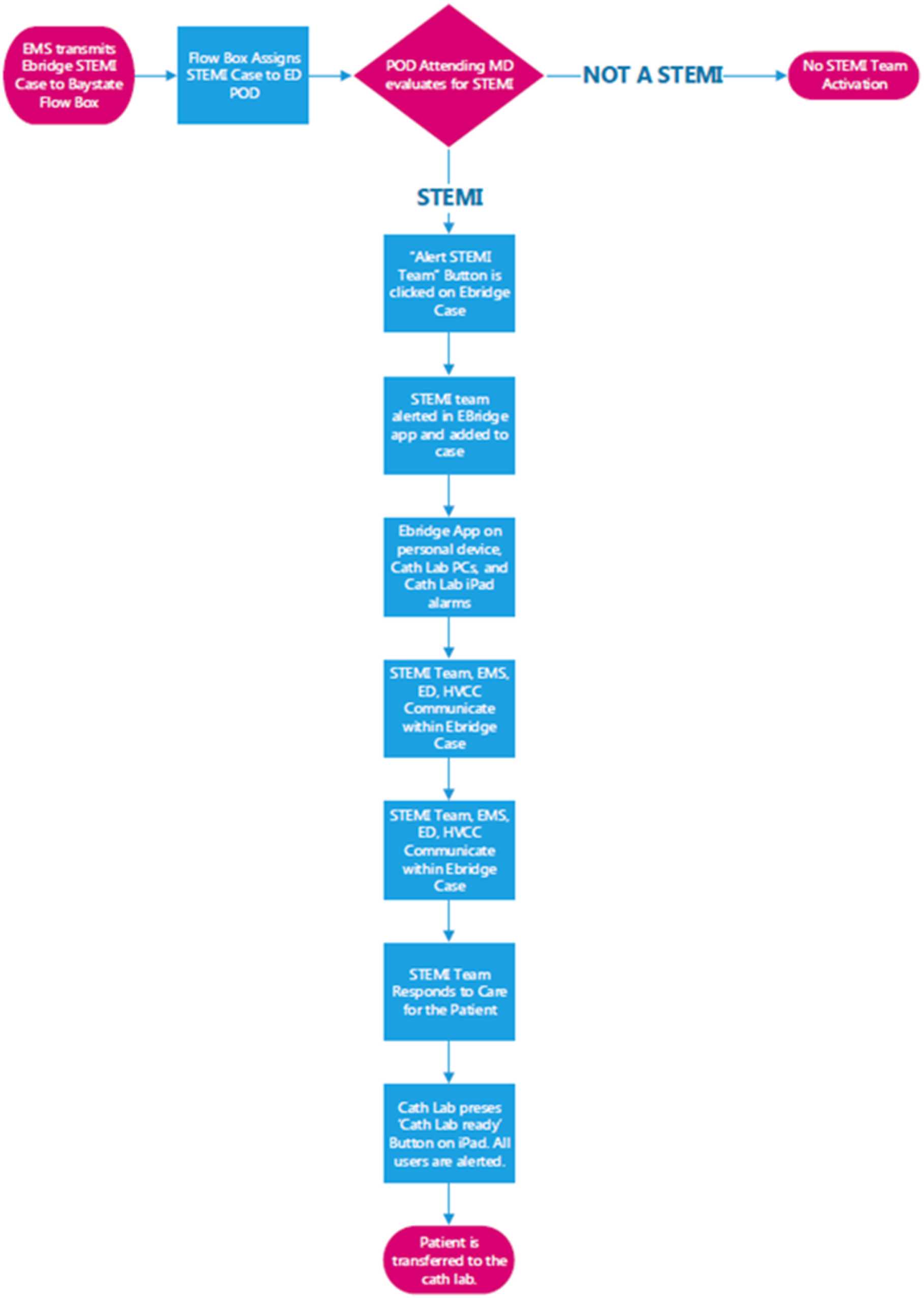

## Results

### Patients

We identified a total of 795 STEMI cases that underwent emergent coronary angiography at our institution between January 2017 and July 2020. After excluding interfacility transfers, cases without indication for or contraindications to PCI, and those who received thrombolytics, our final sample size was 428. Out of these, EMS transmissions using the mobile application were used in 132 patient cases since the application was implemented on May 14th.

When comparing the mean door-to-balloon time, patients who utilized the application had an average time of 61.2 minutes, while those who did not use the application had an average time of 68.4 minutes. This resulted in a mean difference of 7.2 minutes (95% CI -12.2 to 2.14) (Table 1). Additionally, the utilization of the application led to a statistically significant reduction in ED leave time by 4.4 minutes. There was also a trend towards improvement in the first medical contact to balloon time, although it was not statistically significant. However, there was no significant difference in travel time between the two groups.

**Table 1:** Effect of Mobile Application

|  | Mean Time with use of Application (SD) | Mean Time without use of Application (SD) | Mean Difference in Times (minutes) | P value | 95% CI of the difference |
| --- | --- | --- | --- | --- | --- |
| D2b | 61.2 (24.3) | 68.36 (24.61) | -7.17 | 0.005 | -12.2 to -2.14 |
| FMC 2 B | 94.87 (26.52) | 100.24 (28.03) | -5.37 | 0.071 | -11.21 to 0.46 |
| Time in the ED | 24.98 (15.45) | 29.38(17.18) | -4.41 | 0.011 | -7.81 to -1.00 |
| Travel time | 17.82 (8.94) | 19.66 (10.61) | -1.83 | 0.82 | -3.9 to 0.23 |

\*DTB = Door To Balloon Time
\*\*FMC2B = First Medical Contact to Balloon time.

During the day shift, there were a total of 273 cardiac catheterization lab activations, while during the night shift there were 155. Comparing the day and night shifts, the mean door-to-balloon time, first medical contact to balloon time, and boarding time in the emergency department were all significantly lower during the day shift (Table 4). However, when looking specifically at the utilization of the mobile application, there was no statistically significant reduction in these metrics during the day shift. On the other hand, during the night shift, the application resulted in improvements of 10.7 minutes in door-to-balloon time and 8.9 minutes in ED leave time. The mean difference in first medical contact to balloon time was 9.5 minutes, but this difference was not statistically significant. Furthermore, there was no statistically significant difference in travel time for those who utilized the application.

**Table 2:**
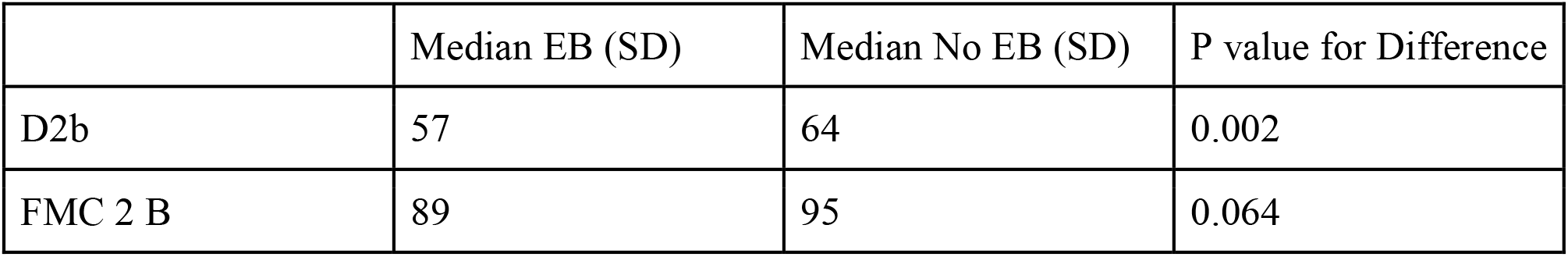
Median Times based on whether Mobile Application was Utilized.

|  | Median EB (SD) | Median No EB (SD) | P value for Difference |
| --- | --- | --- | --- |
| D2b | 57 | 64 | 0.002 |
| FMC 2 B | 89 | 95 | 0.064 |

**Table 3:** Median Test based on whether EBridge was used

|  | Median (minutes) |  | EB | No EB | P value |
| --- | --- | --- | --- | --- | --- |
| D2b | 62 | > Median | 49 | 159 | 0.002 |
|  |  | <= Median | 83 | 137 |  |
| FMC 2 B | 92 | > Median | 56 | 124 | 0.081 |
|  |  | <= Median | 74 | 109 |  |
| Travel Time | 17 | > Median | 58 | 114 | 0.473 |
|  |  | <= Median | 72 | 118 |  |
| Time in the ED | 25 | > Median | 52 | 142 | 0.039 |
|  |  | <= Median | 74 | 127 |  |

EB = E-Bridge Mobile Application.

**Table 4:** Effect of Day Shift and Night Shift on Door to Balloon Time, First Medical Contact to Balloon Time, ED Leave time and Travel time..

|  | Mean Day (SD) | Mean Night (SD) | Mean Difference (minutes) | P value | 95% CI of the difference |
| --- | --- | --- | --- | --- | --- |
| D2b | 63.02 (24.04) | 71.67 (25) | -8.65 | 0.001 | -13.5 to -3.77 |
| FMC 2 B | 94.47 (26.51) | 104.98(28.23) | -10.51 | 0.001 | -16.44 to -4.59 |
| Time in the ED | 24.42 (14.55) | 33.95(18.47) | -9.53 | 0.000 | -13.03 to -6.03 |
| Travel time | 18.94 (10.3) | 19.09 (9.7) | -0.147 | 0.892 | -2.27 - 1.98 |

**Table 5:** Effect of E-Bridge during Day shift (n=273)

|  | Mean w/ EB (SD) | Mean w/o EB (SD) | Mean Difference | P value | 95% CI of the difference |
| --- | --- | --- | --- | --- | --- |
|  |  |  | (minutes) |  |  |
| D2b | 59.78 (22.13) | 64.56 (24.80) | -4.78 | 0.111 | -10.66 to 1.10 |
| FMC 2 B | 92.94 (24.07) | 95.38 (27.90) | -2.43 | 0.486 | -9.30 to 4.44 |
| Time in the ED | 23.67 (14.196) | 24.80 (14.76) | -1.14 | 0.556 | -4.95 to 2.67 |
| Travel time | 17.36 (9.32) | 19.90 (10.77) | -2.54 | 0.062 | -5.21 to 0.13 |

**Table 6:** Effect of EB during Night Shift (n = 155)

|  | Mean w/ EB (SD) | Mean w/o EB (SD) | Mean Difference (minutes) | P value | 95% CI of the difference |
| --- | --- | --- | --- | --- | --- |
| D2b | 64.02 (28.22) | 74.70 (23.04) | -10.68 | 0.029 | -19.23 to -2.019 |
| FMC 2 B | 98.63 (30.73) | 108.11 (26.54) | -9.48 | 0.084 | -20.27 to 1.32 |
| Time in the ED | 27.6 (17.58) | 36.46 (18.29) | -8.87 | 0.008 | -15.32 to -2.41 |
| Travel time | 18.73 (8.15) | 19.27 (10.41) | -0.54 | 0.744 | -3.81 to 2.73 |

## Discussion

The study aimed to evaluate the impact of an app-based communication system on reducing door-to-balloon (DTB) times and first medical contact to balloon (FMC2B) times in patients with ST-Segment Elevation Myocardial Infarctions (STEMI). The results showed a statistically significant reduction in DTB time by 7.2 minutes (p=0.005) and in ED board time by 4.41 minutes (p=0.01) when the app was utilized.

These findings are consistent with the hypothesis that early communication between EMS providers in the field and in-hospital providers can lead to improved STEMI outcomes. By transmitting prehospital history, vitals, and ECGs, the app facilitated more efficient triage, diagnosis, and initial management of STEMI patients, resulting in reduced DTB and ED board times.

It is worth noting that while there was a trend towards improvement in FMC2B time, it did not reach statistical significance. This suggests that the app may have had a greater impact on the in-hospital processes (e.g., activation of the cardiac catheterization lab, transfers, diagnostic clarification) rather than the prehospital phase of care. Further research may be needed to explore strategies specifically targeting FMC2B time reduction.

Additionally, the study analyzed the impact of the app during different shifts and found that the improvements in DTB time and ED leave time were significant during the night shift but not during the day shift. This discrepancy may be attributed to differences in staffing, workflow, or patient volumes between the shifts. Understanding these variations can inform targeted interventions to optimize STEMI care during specific time periods.

The study has several strengths, including a relatively large sample size and a rigorous statistical analysis. However, there are limitations to consider. The study design was retrospective and observational, which may introduce biases and confounding factors. The generalizability of the findings may also be limited to the specific institution and context in which the study was conducted.

In conclusion, the implementation of an app-based communication system between EMS providers and in-hospital providers led to statistically significant reductions in DTB time and ED board time in patients with STEMI. This highlights the potential of technology-enabled communication to improve STEMI care and outcomes. Further research and prospective studies are needed to validate these findings and explore the optimal strategies for implementing such communication systems in different healthcare settings.

## Data Availability

Data available on request from the authorsN

## Abbreviations

D2B: Door to balloon time
EMS: Emergency Medical Services
PCI: Percutaneous Coronary Intervention
STEMI: ST-Segment Elevation Myocardial Infarction
CCL: Cardiac Catheterization Lab

https://pubmed.ncbi.nlm.nih.gov/34323867/

https://pubmed.ncbi.nlm.nih.gov/32938901/

https://pubmed.ncbi.nlm.nih.gov/36712391/

https://pubmed.ncbi.nlm.nih.gov/35851025/

## Notes

### Competing Interest Statement

The authors have declared no competing interest.

### Funding Statement

No external funding was received

### Author Declarations

Study received review and approval for waiver from the Baystate Medical Center Institutional Review Board.

